# JOINT CONTRIBUTION OF MARKERS OF NEURODEGENERATION AND HYPERCHOLESTEROLEMIA TO DEMENTIA RISK

**DOI:** 10.1101/2022.01.16.22269370

**Authors:** Laura Perna, Ute Mons, Hannah Stocker, Léon Beyer, Konrad Beyreuther, Kira Trares, Bernd Holleczek, Ben Schöttker, Robert Perneczky, Klaus Gerwert, Hermann Brenner

## Abstract

**Background:** The examination of markers of neurodegeneration (glial fibrillary acidic protein; GFAP, neurofilament light chain; NfL, phosphorylated tau181; p-tau181) among individuals with high comorbidity of neurodegenerative and cerebrovascular disease and their interplay with vascular risk factors, particularly high cholesterol levels, might contribute to explaining the link between body and brain. The aim of this study was to assess whether the association of GFAP, NfL, and p-tau181 with dementia risk varies depending on levels of total cholesterol (TC) and *APOE* ε4 genotype.

**Methods:** Nested case-control study embedded within a population-based cohort and including 768 older adults (261 dementia cases and 508 randomly selected controls) followed for up to 17 years with regard to clinical diagnosis of various age-related diseases. GFAP, NfL, and p-tau181 were measured in baseline blood samples using the Single-Molecule Array (Simoa) Technology (Quanterix, USA) and categorized into high (quartile 4) versus low (quartiles 1-3). Logistic regression analyses and spline regression models for dose-response analyses were used. ROC curves by cholesterol levels were also calculated.

**Results:** The risk of a dementia diagnosis was significantly increased between participants with high vs. low levels of GFAP and NfL and the risk substantially varied by TC levels. For GFAP and NfL the ORs of a dementia diagnosis were 5.10 (2.45-10.60) and 2.96 (1.43-6.14) in participants with high and 2.44 (1.47-4.07) and 1.15 (0.69-1.92) in those with low TC. *APOE* ε4 genotype further modified the strength of the associations with different patterns, depending on specific marker and type of dementia. No significant association was seen with p-tau181.

**Conclusions:** These results suggest that in the general population blood GFAP and NfL are better predictors of dementia than p-tau181 and that their associations with dementia risk are highly amplified by hypercholesterolemia, also depending on *APOE* ε4 genotype.

## Introduction

Dementia is a progressive disorder characterized by deterioration of cognitive, functional, and behavioral abilities. It has long been established that physical and mental health are fundamentally linked and that a healthy body is an essential precondition of a healthy brain and vice versa. One of the main mechanisms linking body and brain health is the maintenance of a good vascular health^1-5^ and there is strong evidence indicating that several modifiable risk factors for dementia are also risk factors for vascular pathology^6^ and that better cardiovascular health leads to better cognitive health.^2,7-10^

In previous work, we found that the presence of hypercholesterolemia and cardiovascular pathologies modified the association of the apolipoprotein E (*APOE*) ε4 genotype with cognitive function^11^. Here we ask whether blood markers of neurodegeneration contribute to explaining the link between hypercholesterolemia and cognitive decline, also depending on *APOE* ε4 genotype and the presence of vascular pathologies. As blood markers, we selected glial fibrillary acidic protein (GFAP), neurofilament light chain (NfL), and phosphorylated tau181 (p-tau181), because all these markers have been consistently associated with dementia or dementia risk, and GFAP and NfL also with cerebrovascular disease.^12-20^ Altered levels of GFAP, an astrocytic cytoskeletal protein serving as a marker of abnormal activation and proliferation of astrocytes due to neuronal damage, could also point to a possible dysregulation of brain cholesterol synthesized by astrocytes, whose interconnections with peripheral cholesterol are yet unexplored.^21-22^ Furthermore, astrocytes abundantly express ApoE protein, the principal lipid carrier in brain, including cholesterol, and the ε4 allele of the *APOE* gene increases low-density lipoprotein cholesterol,^23^ which is particularly deleterious to vascular health. *APOE* ε4 is also a risk factor for Alzheimer’s disease (AD) and, to a lesser extent, for vascular dementia.^24-25^

Most dementia biomarker studies have been conducted in clinic-based cohorts including very narrow populations^26-27^ and mostly focusing on AD-pathology. We believe that in order to understand the impact of markers of neurodegeneration on dementia fully, we need to focus on representative populations with mixed pathologies (comorbidity of neurodegenerative and cerebrovascular disease) and analyze how such markers unfold in synergism with other risk factors. Here, we aimed to focus on total cholesterol (TC) and investigated the association between GFAP, NfL, and p-tau181 with dementia risk by exploring whether the strength of the association varied depending on levels of TC, *APOE* ε4 genotype, and the presence of vascular pathology prior to or at the time of the measurement of markers of neurodegeneration.

## Methods

### Study population

Data are based on a nested case-control study within a community-based prospective cohort of older adults followed for up to 17 years with regard to clinical diagnosis of various age-related diseases and mortality (the ESTHER study). ESTHER participants (n=9,940) were recruited in 2000-2002 in Saarland, a southwestern German state. Eligibility criteria were age between 50-75 years, sufficient knowledge of the German language, residence in Saarland, and willingness to attend a general health examination performed by general practitioners (GPs).^28^ No specific exclusion criteria were applied, as this would have impaired the generalizability of the ESTHER study.^29^ Both at baseline and at follow-up measurements, participants provided health information and biological samples, including blood samples, which were stored at -80°C. GPs provided available medical records. The ESTHER study was approved by the Ethics Committee of the Medical Faculty at Heidelberg University and the Physicians’ Board of Saarland. All participants provided written informed consent.

### Dementia assessment

During the 14- and 17-year follow-up, the GPs of 8,353 ESTHER study participants (84%) could still be contacted and were asked to provide information relating to a diagnosis of dementia since enrollment and to send the corresponding medical records, if available. GPs of participants who had dropped out of the study or had died were also contacted. In total, GPs of n=6,357 participants provided dementia diagnosis information. Dementia diagnoses were made in community settings by different medical doctors according to the International Classification of Diseases (ICD-10), the official classification for the encoding of medical diagnoses in Germany, and with heterogeneous diagnostic procedures as could be inferred from the available medical records.

The present dataset included 507 controls (participants that remained without dementia diagnosis throughout follow-up) chosen at random and 261 dementia cases (dementia diagnosis occurring between baseline and the 17-year follow-up), as previously described.^30^ Among the dementia cases there were n=163 participants with a diagnosis of the clinical syndrome of AD (AD dementia), thereof n=18 with an additional diagnosis of vascular dementia (VaD) or mixed dementia, and n=83 with a diagnosis of VaD, thereof 17 with an additional diagnosis of AD dementia or mixed dementia. For n=15 only a diagnosis of mixed dementia was reported. However, a close examination of the available medical records revealed that comorbidity of neurodegenerative and cerebrovascular pathologies were highly prevalent among all dementia cases. In light of results of clinical-pathological studies performed among population-based cohorts showing that vascular pathologies are the most prevalent pathologies among demented patients,^31-32^ and based on the observation that pure AD pathology is very rare,^33^ we hence assume that in the ESTHER cohort even in cases where a specific dementia type is reported as primary diagnosis, mixed pathologies were likely to be present in the great majority of dementia cases. In the following, we will use the term dementia risk to indicate the risk of a clinical diagnosis of dementia reflecting a composite endpoint including AD, VaD, and mixed dementia, with AD dementia and VaD indicating predominance of clinical symptoms closer to neurodegenerative or vascular pathologies, respectively.

### Laboratory measurements

#### Markers of neurodegeneration

GFAP, NfL, and p-tau181 were measured in a single batch in lithium-heparin plasma of baseline samples at the Center for Protein Diagnostics (PRODI) of Bochum University (Germany) using the single molecule array (Simoa) Neurology 4-Plex E Advantage Kit and Simoa pTau-181 Advantage V2 Kit (Quanterix, USA) on a HD-X Analyzer as per manufacturer’s instructions. A detailed description of the measurements can be found in a previous work of our group.^30^ All three markers were grouped in a joint category related to neurodegeneration because their elevation is indicative of underlying neurodegenerative dementias.

#### Total cholesterol

Cholesterol levels were determined in all ESTHER participants at baseline, with samples taken at the same time as the plasma used to measure markers of neurodegeneration. Cholesterol concentrations were measured in serum using a timed-endpoint method by which cholesterol esterase hydrolyzes esters to free cholesterol (Beckman Coulter SYNCHRON System(s)). The measurements were performed in the Labor of the University Clinic Heidelberg, Germany. Hypercholesterolemia was defined as TC ≥ 240 mg/dL.^34-35^

#### APOE genotype

εalleles were determined based on allelic combinations of single nucleotide polymorphisms (SNP) rs7412 and rs429358 using TaqMan SNP genotyping assays. Genotypes were analyzed in an endpoint allelic discrimination read using a PRISM 7000 Sequence detection system (Applied Biosystems). In some cases with missing values (n= 38) the *APOE* genotype could be determined based on *APOE* SNP results from GWAS data. Participants were divided into carriers of the ε4 allele (ε2/ε4, ε3/ε4, ε4/ε4) and non-carriers (ε2/ε2, ε2/ε3, ε3/ε3).

### Cardiovascular diseases (CVD) and sociodemographic data

CVD used for the analyses were diagnosed either at or prior to baseline and defined as combined category including stroke, myocardial infarction, coronary artery disease, or coronary revascularization. Age, sex, and educational level (less, equal to, or higher than 9 years of school education) were collected through the self-administered questionnaire administered at baseline.

### Statistical analysis

Main baseline characteristics of the included ESTHER cases and controls were described for the whole population and according to levels of TC. Multivariate logistic regression models adjusted for age, sex, educational level, TC, and *APOE* ε4 genotype were estimated for the outcomes dementia, AD dementia, and VaD. Values of markers of neurodegeneration were divided into quartiles (Q), and odds ratios (OR) with 95% confidence intervals (CI) were calculated for the highest quartile (Q4) compared with the other three quartiles (Q1-3), serving as the reference group. All regression models were run for the overall sample and, separately, by levels of TC (high: ≥ 240 mg/dL; low < 240 mg/dL).^34-35^ Additional regression models adjusted for age, sex, educational level, and TC were performed stratified by *APOE* ε4 genotype (carriers/non-carriers) and by CVD. To test for significant differences in associations of the biomarkers with dementia risk between TC and CVD subgroups, interaction terms for the respective biomarker with TC or CVD, respectively, were added to all models.

Dose-response analyses using restricted cubic spline functions (RCS) with knots at the 5^th^, 35^th^, 65^th^, and 95^th^ percentiles were conducted. A receiver operating characteristic (ROC) curve was calculated in order to explore the discriminative power of the markers of neurodegeneration in addition to age, sex, educational level, and *APOE* ε4. RCS functions and ROC curves were derived separately in subgroups with high and low TC; the values of GFAP, NfL, and p-tau181 were inserted as continuous variables. For all three biomarkers, the distributions were moderately right skewed. However, since the data were reasonably close to a normal distribution and for better interpretability, we used the original values but performed sensitivity analyses with log-transformed values. All analyses were performed with the statistical software SAS®, version 9.4, Cary, NC, USA.

## Results

In this nested case-control study, the majority of cases and controls was older than sixty years and a large majority had a low educational level (**Table 1**). In total, n=94 (36%) cases and n=194 (38%) controls had high TC and both among cases and controls this percentage was higher among *APOE* ε4 carriers than among non-carriers (cases: 42% vs. 32%, controls: 41% vs. 38%). For all three markers of neurodegeneration, levels were higher among cases.

**Table 1.** Main characteristics of the study population.

| ESTHER study<br>(n=768) | Cases |  |  | Controls |  |  |
| --- | --- | --- | --- | --- | --- | --- |
|  | overall<br>n=261 | high TC <sup>a</sup><br>n=94 | low TC <sup>b</sup><br>n=167 | overall<br>n=507 | high TC <sup>a</sup><br>n=194 | low TC <sup>b</sup><br>n=312 |
|  | n (%) | n (%) | n (%) | n (%) | n (%) | n (%) |
| **Age groups** |  |  |  |  |  |  |
| 50-59 | 20 (7.7) | 8 (8.5) | 12 (7.2) | 202 (39.8) | 79 (40.7) | 123 (39.4) |
| 60-70 | 152 (58.2) | 54 (57.5) | 98 (58.7) | 258 (50.9) | 101 (52.1) | 156 (50.0) |
| 71-75 | 89 (34.1) | 32 (34.0) | 57 (34.1) | 47 (9.3) | 14 (7.2) | 33 (10.6) |
| **Sex** |  |  |  |  |  |  |
| women | 151 (57.9) | 64 (68.1) | 87 (52.1) | 278 (54.8) | 120 (61.9) | 158 (50.6) |
| men | 110 (42.1) | 30 (31.9) | 80 (47.9) | 229 (45.2) | 74 (38.1) | 154 (49.4) |
| **Educational level** |  |  |  |  |  |  |
| low <sup>c</sup> | 214 (82.0) | 75 (79.8) | 139 (83.2) | 394 (77.7) | 156 (80.4) | 238 (76.3) |
| high <sup>d</sup> | 37 (14.2) | 15 (16.0) | 22 (13.2) | 102 (20.1) | 34 (17.5) | 67 (21.5) |
| missing values | 10 (3.8) | 4 (4.3) | 6 (3.6) | 11 (2.2) | 4 (2.1) | 7 (2.2) |
| **APOE ε4** |  |  |  |  |  |  |
| carriers <sup>e</sup> | 103 (39.5) | 43 (45.7) | 60 (35.9) | 131 (25.8) | 54 (27.8) | 77 (24.7) |
| non-carriers <sup>f</sup> | 143 (54.8) | 46 (48.9) | 97 (58.1) | 364 (71.8) | 137 (70.6) | 226 (72.4) |
| missing values | 15 (5.8) | 5 (5.3) | 10 (6.0) | 12 (2.4) | 3 (1.6) | 9 (2.9) |
| **CVD <sup>g</sup>** |  |  |  |  |  |  |
| yes | 76 (29.1) | 19 (20.2) | 57 (34.1) | 101 (19.9) | 32 (16.5) | 69 (22.1) |
| no | 185 (70.9) | 75 (79.8) | 110 (65.9) | 406 (80.1) | 162 (83.5) | 243 (77.9) |
|  | **mean (SD)** | **mean (SD)** | **mean (SD)** | **mean (SD)** | **mean (SD)** | **Mean (SD)** |
| **GFAP** | 133.2 (78.2) | 146.0 (99.1) | 126.0 (62.6) | 87.0 (46.7) | 83.4 (40.8) | 89.3 (50.0) |
| **NfL** | 22.8 (13.4) | 24.1 (14.8) | 22.1 (12.6) | 15.8 (8.4) | 15.8 (9.9) | 15.8 (7.4) |
| **P-tau181** | 2.1 (1.5) | 2.3 (1.5) | 2.1 (1.5) | 1.7 (1.2) | 1.7 (1.5) | 1.7 (1.0) |

TC, total cholesterol; SD, standard deviation; *APOE*, Apolipoprotein E; GFAP, Glial Fibrillary Acidic Protein; NfL, Neurofilament light; p-tau181, tau phosphorylated at threonine 181.
a) TC ≥ 240 mg/dL
b) TC < 240 mg/dL
c) ≤ 9 years of school education
d) > 9 years of school education
e) participants carrying *APOE* ε4 genotype ( ε2 ε4, ε3 ε4, ε4 ε4)
f) participants not carrying *APOE* ε4 genotype ( ε2 ε2, ε3 ε2, ε3 ε3)
g) CVD was defined as combined endpoint of stroke, or myocardial infarction, or coronary artery disease, or revascularization of coronary arteries, or pulmonary embolism.

Results of regression analyses showed that the odds of a dementia diagnosis were much higher among participants with both high baseline levels of markers of neurodegeneration and high TC levels than among participants with low TC (**Table 2**). This pattern was particularly evident for GFAP and NfL, for which ORs among participants with high TC were 5.10 (CI 2.45-10.60) and 2.96 (CI 1.43-6.14), respectively. For comparison, the ORs among participants with low TC were 2.44 (CI 1.47-4.07) and 1.15 (CI 0.69-1.92), respectively. Models including interaction terms yielded a statistically significant interaction between NfL and TC with respect to dementia risk (p=0.037).

**Table 2.** Associations of high^a^ GFAP, NfL, and p-tau181 with dementia risk by total cholesterol and *APOE* ε4 status (ESTHER cohort study 2000-2017)

|  | Overall | High TC <sup>b</sup> | Low TC <sup>c</sup> |
| --- | --- | --- | --- |
| *General population*<br>ESTHER study<br>(n=768) | **Odds ratio**<br>(95%CI;<br>n= cases) | **Odds ratio**<br>(95%CI;<br>n= cases) | **Odds ratio**<br>(95%CI;<br>n= cases) |
| **Glial Fibrillary Acidic Protein** |  |  |  |
| Overall | 3.08<br>(2.04-4.66)<br>(n=261) | 5.10<br>(2.45-10.60)<br>(n=94) | 2.44<br>(1.47-4.07)<br>(n=167) |
| APOE ε4+<sup>d</sup> | 3.13<br>(1.63-6.04)<br>(n=103) | 2.40<br>(0.82-7.01)<br>(n=43) | 3.90<br>(1.67-9.13)<br>(n=60) |
| APOE ε4-<sup>e</sup> | 3.41<br>(1.98-5.88)<br>(n=143) | 10.02<br>(3.57-28.15)<br>(n=46) | 2.05<br>(1.06-3.97)<br>(n=97) |
| **Neurofilament light** |  |  |  |
| Overall | 1.57<br>(1.04-2.38)<br>(n=261) | 2.96<br>(1.43-6.14)<br>(n=94) | 1.15<br>(0.69-1.92)<br>(n=167) |
| APOE ε4+<sup>d</sup> | 1.30<br>(0.62-2.70)<br>(n=103) | 4.21<br>(1.15-15.40)<br>(n=43) | 0.67<br>(0.26-1.74)<br>(n=60) |
| APOE ε4-<sup>e</sup> | 1.82<br>(1.10-3.01)<br>(n=143) | 2.42<br>(0.98-5.97)<br>(n=46) | 1.55<br>(0.84-2.85)<br>(97) |
| **p-tau181** |  |  |  |
| Overall | 1.29<br>(0.86-1.93)<br>(n=261) | 1.51<br>(0.77-2.98)<br>(n=94) | 1.19<br>(0.72-1.97)<br>(n=167) |
| APOE ε4+<sup>d</sup> | 0.99<br>(0.52-1.91)<br>(n=103) | 1.10<br>(0.38-3.18)<br>(n=43) | 0.97<br>(0.41-2.26)<br>(n=60) |
| APOE ε4-<sup>e</sup> | 1.67<br>(1.00-2.79)<br>(n=143) | 2.02<br>(0.83-4.94)<br>(n=46) | 1.55<br>(0.82-2.93)<br>(n=97) |

GFAP, Glial Fibrillary Acidic Protein; NfL, Neurofilament light; p-tau181, tau phosphorylated at threonine 181; APOE, Apolipoprotein E; TC, total cholesterol; CI, confidence interval.
a) ≥ quartile 4 (vs quartiles 1-3)
b) TC ≥ 240 mg/dL
c) TC < 240 mg/dL
d) participants carrying APOE ε4 genotype (ε2ε4, ε3ε4, ε4ε4)
e) participants not carrying APOE ε4 genotype (ε2ε2, ε3ε2, ε3ε3)
All models were adjusted for age, sex, educational level, APOE ε4 genotype, and TC. Subgroup analyses by TC were adjusted for age, sex, educational level, and APOE ε4. Subgroup analyses by TC and APOE ε4 genotype were adjusted for age, sex, and educational level.

The additional stratification of the models by *APOE* ε4 genotype revealed particularly high odds of dementia, with an OR of 10.02 (CI 3.57-28.15) among *APOE* ε4 non-carriers with high GFAP and high TC, and an OR of 4.21 (CI 1.15-15.40) among carriers with high NfL and high TC. While the interaction between GFAP and TC was statistically significant in the *APOE* ε4-subgroup (p=0.012), the interaction between NfL and TC was statistically significant in the *APOE* ε4+ subgroup (p= 0.018). Interactions including p-tau181 were not statistically significant.

High levels of GFAP at baseline were also associated with 12-fold odds of dementia among participants with CVD, compared to roughly twofold odds among those free of CVD at baseline (**Table 3**). A similar pattern could not be observed for participants with high levels of NfL or p-tau181. By contrast, high levels of NfL and p-tau181 were statistically significantly associated with dementia in the group without baseline CVD. Adding interaction terms to the models yielded statistically significant interactions between CVD and GFAP (p=0.010) and between CVD and p-tau181 (p=0.040). In regression models not including markers of neurodegeneration and adjusted for age, sex, educational level, and *APOE* ε4, neither hypercholesteremia nor prevalence of CVD at baseline were independently associated with increased odds of dementia (OR 0.95; CI 0.66-1.37 and OR 1.04, CI 0.69-1.57, respectively).

**Table 3.** Associations of high^a^ GFAP, NfL, and p-tau181 with dementia risk among participants with and without CVD (ESTHER cohort study 2000-2017)

|  | Glial Fibrillary<br>Acidic Protein | Neurofilament<br>light | P-tau181 |
| --- | --- | --- | --- |
| ESTHER study<br>(n=768) | Odds ratio<br>(95%CI) | Odds ratio<br>(95%CI) | Odds ratio<br>(95%CI) |
| **Subgroup with CVD<sup>b</sup>**<br>(n=177) |  |  |  |
| Dementia<br>(n= 76) | 12.47<br>(4.43-35.06) | 1.26<br>(0.59-2.69) | 0.72<br>(0.31-1.66) |
| Vascular dementia<br>(n= 31) | 6.12<br>(2.10-17.82) | 1.41<br>(0.54-3.67) | 1.64<br>(0.60-4.50) |
| Alzheimer's clinical<br>syndrome<br>(n= 39) | 3.63<br>(1.45-9.13) | 0.80<br>(0.32-1.96) | 0.68<br>(0.26-1.76) |
| **Subgroup without CVD<sup>b</sup>**<br>(n=591) |  |  |  |
| Dementia<br>(n= 185) | 2.22<br>(1.37-3.58) | 1.81<br>(1.10-2.98) | 1.64<br>(1.03-2.62) |
| Vascular dementia<br>(n= 52) | 1.80<br>(0.90-3.58) | 2.04<br>(1.02-4.06) | 1.95<br>(1.01-3.76) |
| Alzheimer's clinical<br>syndrome<br>(n= 124) | 1.78<br>(1.07-2.94) | 1.34<br>(0.79-2.28) | 1.54<br>(0.94-2.52) |

GFAP, Glial Fibrillary Acidic Protein; NfL, Neurofilament light; p-tau181, tau phosphorylated at threonine 181; CVD, cardiovascular disease; CI, confidence interval.
a) ≥ quartile 4 (vs quartiles 1-3)
b) CVD was defined as combined endpoint of stroke, or myocardial infarction, or coronary artery disease, or revascularization of coronary arteries, or pulmonary embolism.
All models were adjusted for age, sex, educational level, *APOE* ε4 genotype, and total cholesterol.

Regarding the type of dementia for GFAP the odds for VaD were higher among *APOE* ε4 carriers than among non-carriers, while the odds for AD dementia were higher among non-carriers than carriers, particularly in the presence of high TC (**Supplemental Table 1**). For NfL, an opposite pattern could be observed in the presence of high TC (**Supplemental Table 2**). For p-tau181, associations with VaD were stronger among *APOE* ε4 carriers than among non-carriers and in the presence of high TC. (**Supplemental Table 3**).

Restricted cubic spline curves indicated a steadily increasing dose-response association between higher GFAP levels and dementia among participants with TC ≥240mg/dL (**Figure 1**). For the other markers, dementia risk seemed to increase more strongly in participants with high TC levels, but leveled off with higher biomarker levels or even showed a tendency to decrease at higher NfL levels. ROC curve analyses for dementia also showed that GFAP was the marker with the strongest discriminative ability in this cohort. Specifically, the area under the ROC curve (AUC) increased from 0.811 (95%CI 0.758-0.864) for the main model to 0.841 (0.791-0.891) after the inclusion of GFAP in the group of participants with high TC and from 0.768 (95%CI 0.724-0.813) to 0.780 (95%CI 0.737-0.824) in the group with low TC (**Figure 2**). The inclusion of NfL and p-tau181 to the main model increased the AUC only marginally. Sensitivity analyses performed with the log transformed values yielded results largely similar to those obtained with the original values.

**Figure 1.**
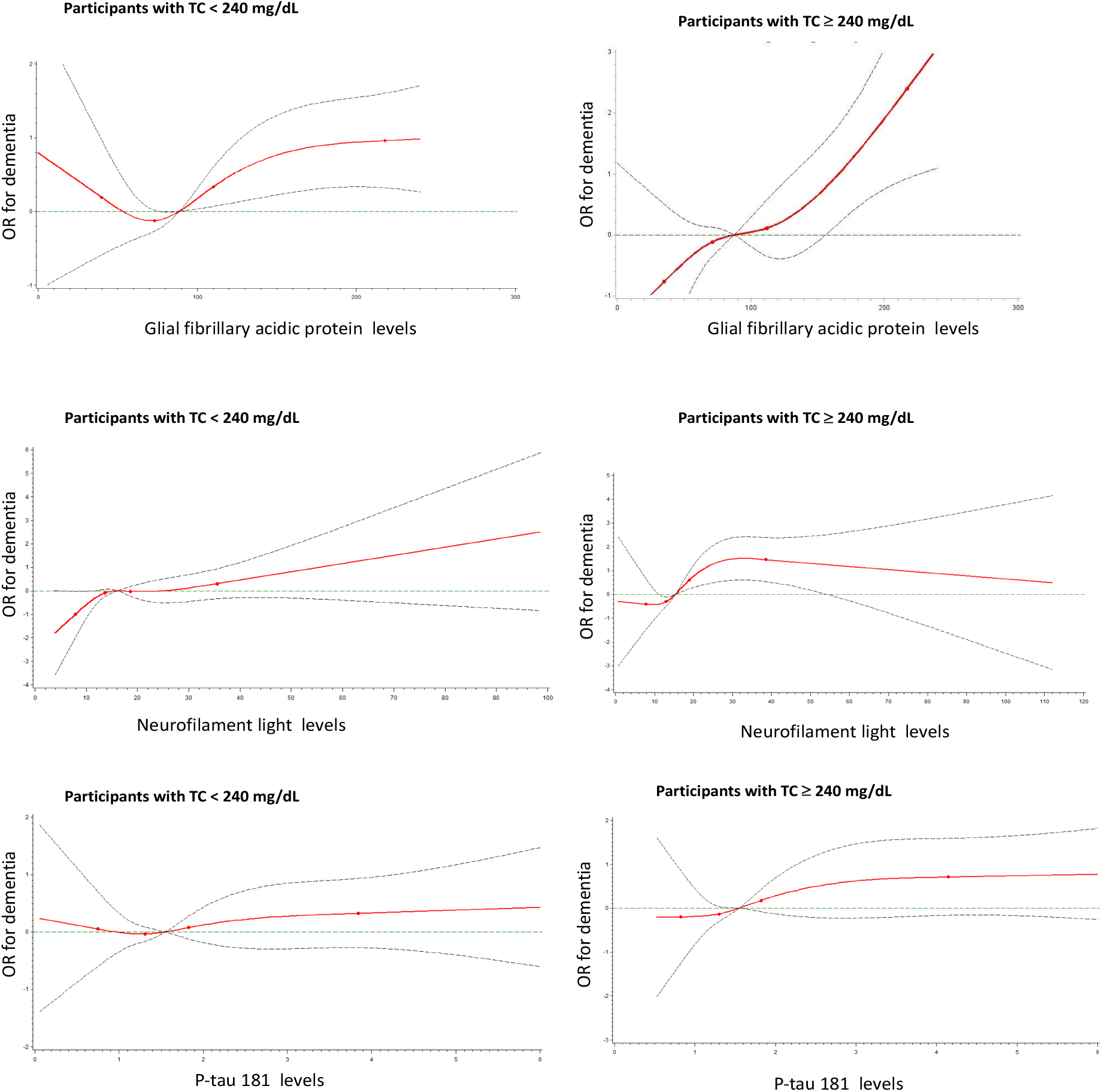
Dose-response associations of dementia by levels of total cholesterol; results of restricted cubic spline regression models. *Top row:* Association of dementia with GFAP (Glial Fibrillary Acidic Protein) among participants with low (< 240 mg/dL) and high (**≥** 240 mg/dL) total cholesterol (TC) levels. *Middle row:* Association of dementia with NfL (Neurofilament light) among participants with low (< 240 mg/dL) and high (**≥** 240 mg/dL) TC levels. *Bottom row:* Association of dementia with p-tau 181 (tau phosphorylated at threonine 181) among participants with low (< 240 mg/dL) and high (**≥** 240 mg/dL) TC levels. Results of spline regression models adjusted for age, sex, educational level, and *APOE* ε4 polymorphism (ε2ε4, ε3ε4, ε4ε4 vs ε2ε2, ε3ε2, ε3ε3). GFAP, NfL, and p-tau 181 were inserted in the models as continuous variables. Solid line: point estimates; dashed curved lines: 95% confidence interval limits; dashed horizontal line: reference line (odds ratio =1); dots: knots (5^th^, 35^th^, 65^th^, and 95^th^).

**Figure 2.**
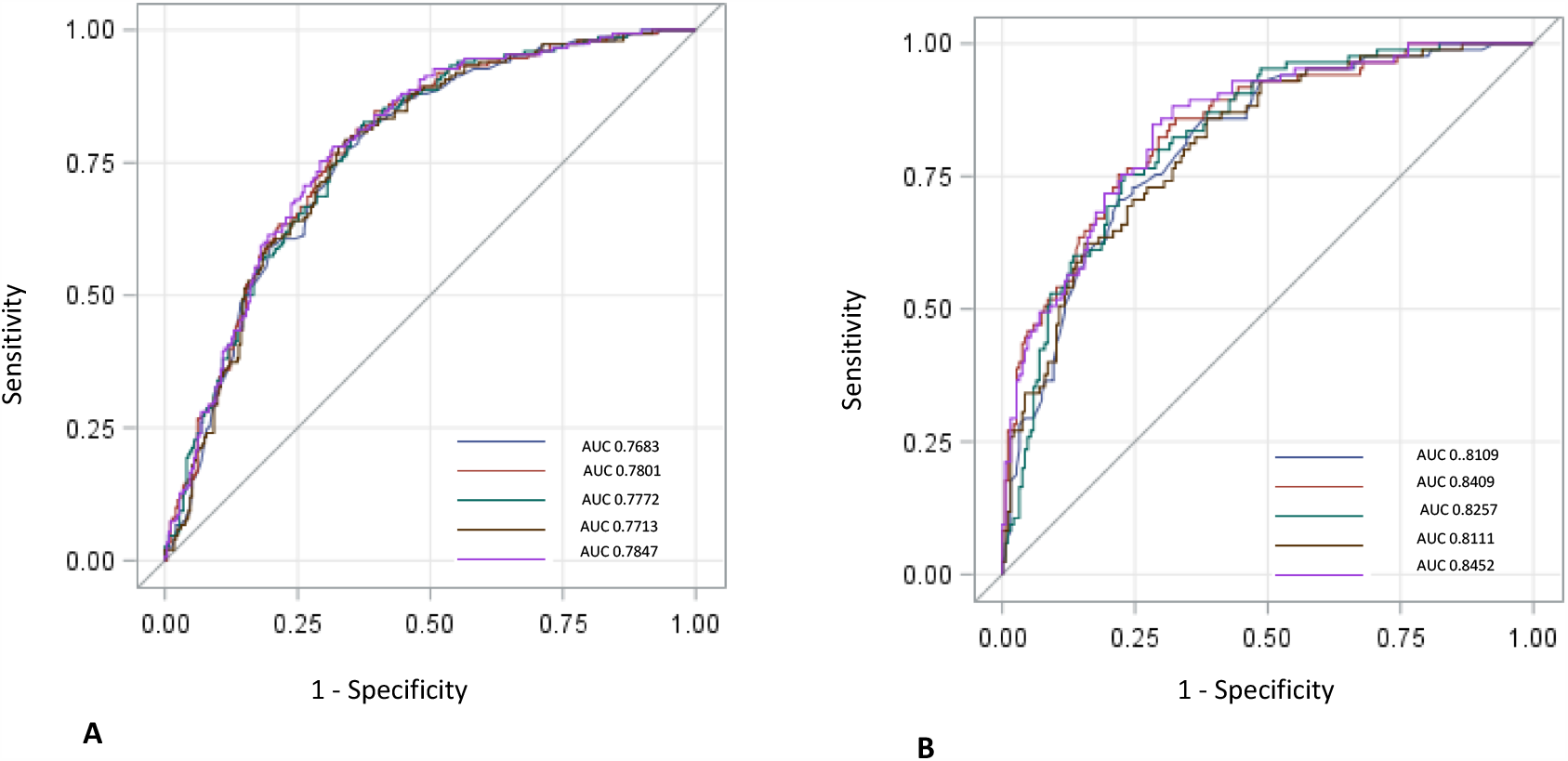
Results of ROC curve analyses for dementia by levels of total cholesterol. **A** Receiver operating characteristics (ROC) curve analyses for dementia with values of the corresponding areas under the curve (AUC) among participants with total cholesterol levels **<** 240 mg/dL. The main model includes age, sex, educational level, and *APOE* ε4 polymorphism (ε2ε4, ε3ε4, ε4ε4 vs ε2ε2, ε3ε2, ε3ε3). Glial Fibrillary Acidic Protein (GFAP), Neurofilament light (NfL), and tau phosphorylated at threonine 181 (P-tau 181) were inserted in the models as continuous variables. 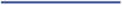 Main model; 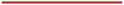 Main model + GFAP; 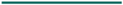 Main model + NfL; 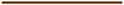 Main model + P-tau 181; 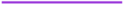 Main model + GFAP, NfL, and P-tau 181. **B** Receiver operating characteristics (ROC) curve analyses for dementia with values of the corresponding areas under the curve (AUC) among participants with total cholesterol levels **≥** 240 mg/dL. The main model includes age, sex, educational level, and *APOE* ε4 polymorphism (ε2ε4, ε3ε4, ε4ε4 vs ε2ε2, ε3ε2, ε3ε3). Glial Fibrillary Acidic Protein (GFAP), Neurofilament light (NfL), and tau phosphorylated at threonine 181 (P-tau 181) were inserted in the models as continuous variables. 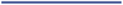 Main model; 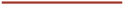 Main model + GFAP; 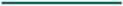 Main model + NfL; 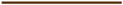 Main model + P-tau 181; 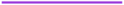 Main model + GFAP, NfL, and P-tau 181.

## Discussion

In this study, we examined associations of markers of neurodegeneration with dementia risk in a population with a high prevalence of mixed pathology and found that the strength of the associations dramatically changed depending on levels of TC and on *APOE* ε4 genotype. We also found that the association of GFAP, but not of NfL and p-tau181, with dementia risk was strongly augmented among people with CVD, independently of hypercholesterolemia. Additionally, the findings showed that GFAP and, to a lesser extent, NfL are more promising than p-tau181 for predicting dementia risk in the older general population with high cardiovascular burden.

Since hypercholesterolemia and *APOE* ε4 are crucial for cardiovascular health,^23,35^ these findings strongly support a vascular link between brain and body health. However, they also suggest that high TC and CVD as risk factors for dementia might become evident only in synergism with other markers or pathologies and, possibly, with genetic information relating to *APOE* ε4 status. This observation would explain the inconsistent findings reported in the literature relating to associations of hypercholesterolemia, CVD, and dementia.^36-38^

The link between vascular pathology and dementia is supported by the relationship between neural activity and cerebral blood flow (neurovascular coupling).^3,39-40^ Neurovascular coupling is a multidimensional process involving several agents and signals,^3^ with astrocytes playing an instrumental role.^41-42^ This might contribute to explaining why GFAP, a marker of activated astrocytes, was the strongest predictor of dementia in this cohort and why the strength of the association of GFAP with dementia risk was mostly increased in an unhealthy vascular milieu.

Furthermore, although brain and peripheral cholesterol are separated by the blood-brain barrier (BBB) and are regulated independently, they can be interconnected and both brain and peripheral cholesterol dysfunction can contribute to dementia.^21,43^ Hence, high GFAP levels might also point to a possible dysregulation in brain cholesterol produced by astrocytes, which might have synergistic effects with peripheral cholesterol for dementia development. Furthermore astrocytes also contribute to the maintenance of the BBB and high GFAP levels might indicate a malfunctioning of the BBB.

In a recent study, GFAP was not associated with *APOE* ε4, but GFAP levels were highest among *APOE* ε4 non-carriers with AD pathology.^13^ The focus on TC in this study allowed us to expand those observations and show that dementia and AD dementia risk was much higher in non-carriers with high TC than in carriers, pointing to a possible synergism between high GFAP and non-carrier status for dementia and AD dementia. For VaD, an opposite pattern could be observed, but due to the small number of participants with VaD, the results relating to this subgroup should be interpreted with caution.

NfL is a blood marker of axonal damage and has been found elevated both in dementia, including AD, and in cerebrovascular disease^20,44^, and high NfL levels have been associated with an increased risk of all-cause dementia, AD, and AD progression.^12,44-45^ Furthermore, it has been shown that an elevated vascular risk factor burden might synergistically interact with AD pathophysiology contributing to longitudinal increases in plasma NfL and cognitive decline.^46^ Our results expand these previous observations by pointing to NfL as a marker for dementia with mixed pathology, especially among *APOE* ε4 carriers with hypercholesterolemia.

Blood p-tau181 has been shown to be a marker associated with progressive AD-related neurodegeneration and capable of distinguishing AD from VaD and other neurodegenerative disorders in previous research.^12,16^ These results have been found in well-characterized cohorts with evidence of AD pathology in the brain. In our community-based cohort including participants with mixed pathology, p-tau181 was weakly associated with dementia and the results were not statistically significant. The presence of hypercholesterolemia had a marginal impact on the strength of the association, except for VaD, which might support previous observations showing associations of accumulation of tau pathology with vascular cognitive impairment.^47^ Our findings point to the need to further examine the role of vascular pathologies and vascular risk factors in the association between p-tau181 with dementia.

The findings of the present study strongly support results of our previous study showing that hypercholesterolemia and CVD changed the association of *APOE* ε4 with cognitive function in two independent cohorts^11^ and provide observational evidence to clinicopathological data from population-based cohorts suggesting an incremental risk of dementia in patients with concomitant vascular and neurodegenerative pathologies compared with patients with only AD-type pathology.^31^ Our findings contribute to explaining this increased dementia risk by suggesting a mechanism involving genetic predisposition and a crosstalk between central and peripheral cholesterol metabolism, possibly driven by the permeabilization of the BBB.^21^ Several factors would contribute to permeabilization of the BBB in this cohort and they include micro- and macroangiopathies, vascular risk factors, inflammatory processes driven by high peripheral cholesterol, and astrocyte damage. In turn, the permeabilization of the BBB could facilitate the release of GFAP in the peripheral circulation.^48^ Another contributing factor for the increased dementia risk is the possible over-production of astrocyte-derived cholesterol driven by abnormal activation and proliferation of astrocytes, which in turn leads to increased AD-like pathology, as shown in lab mice.^48^ These mechanisms involving CVD and dysregulation of central and peripheral cholesterol would also explain why high GFAP levels as a marker for dementia works exceptionally well in this sample with mixed pathologies and has a stronger association with VaD than with AD dementia in this unselected population. The complexity and the multifariousness of the role of cholesterol and CVD in dementia also contribute to explaining the inconsistent finding in the literature relating to CVD and hypercholesterolemia for dementia and suggest that blood levels of GFAP could serve both for assessment of dementia risk and for evaluating effects of dementia prevention strategies addressing hypercholesterolemia. Future studies performed both in human cohorts and in vitro will be crucial in understanding the biological mechanisms underpinning these novel observations.

The coeval measurements of TC and markers of neurodegeneration prevented a temporal exploration of the observed association and limited its interpretation. Other important limitations were the lack of brain biomarkers for AD-pathology and the lack of standardization of dementia diagnoses. Nevertheless, the use of a population-based cohort with 17-year follow-up and high comorbidity of neurodegenerative and cerebrovascular disease reflecting the majority of dementia cases in community settings allowed the identification of GFAP, and to a less extent NfL, as very strong markers for dementia, especially in a milieu with high circulating peripheral cholesterol.

Further studies shall expand such observations to subgroups with low- and high-density lipoprotein cholesterol in combination with other vascular risk factors, especially diabetes mellitus and hypertension and shall especially focus on comprehensive analyses relating to GFAP and mixed dementia in unselected cohorts.

## Data Availability

Due to restrictions of informed consent of the ESTHER study, we are not allowed making the dataset publicly available. However, all data produced in the present study are available upon reasonable request to the authors.

## Finding/support and role of funder/sponsor

The ESTHER study was funded by grants from the Saarland State Ministry for Social Affairs, Health, Women and Family Affairs (Saarbrücken, Germany), the Baden-Württemberg State Ministry of Science, Research and Arts (Stuttgart, Germany), the Federal Ministry of Education and Research (Berlin, Germany) and the Federal Ministry of Family Affairs, Senior Citizens, Women and Youth (Berlin, Germany). UM is supported by the Marga and Walter Boll Foundation, Kerpen, Germany.

None of the funding sources had a role in the conduct of the study, in the analysis or interpretation of the data, in the writing of this manuscript or in the decision to submit it for publication.

---

**Supplemental Table 1.**
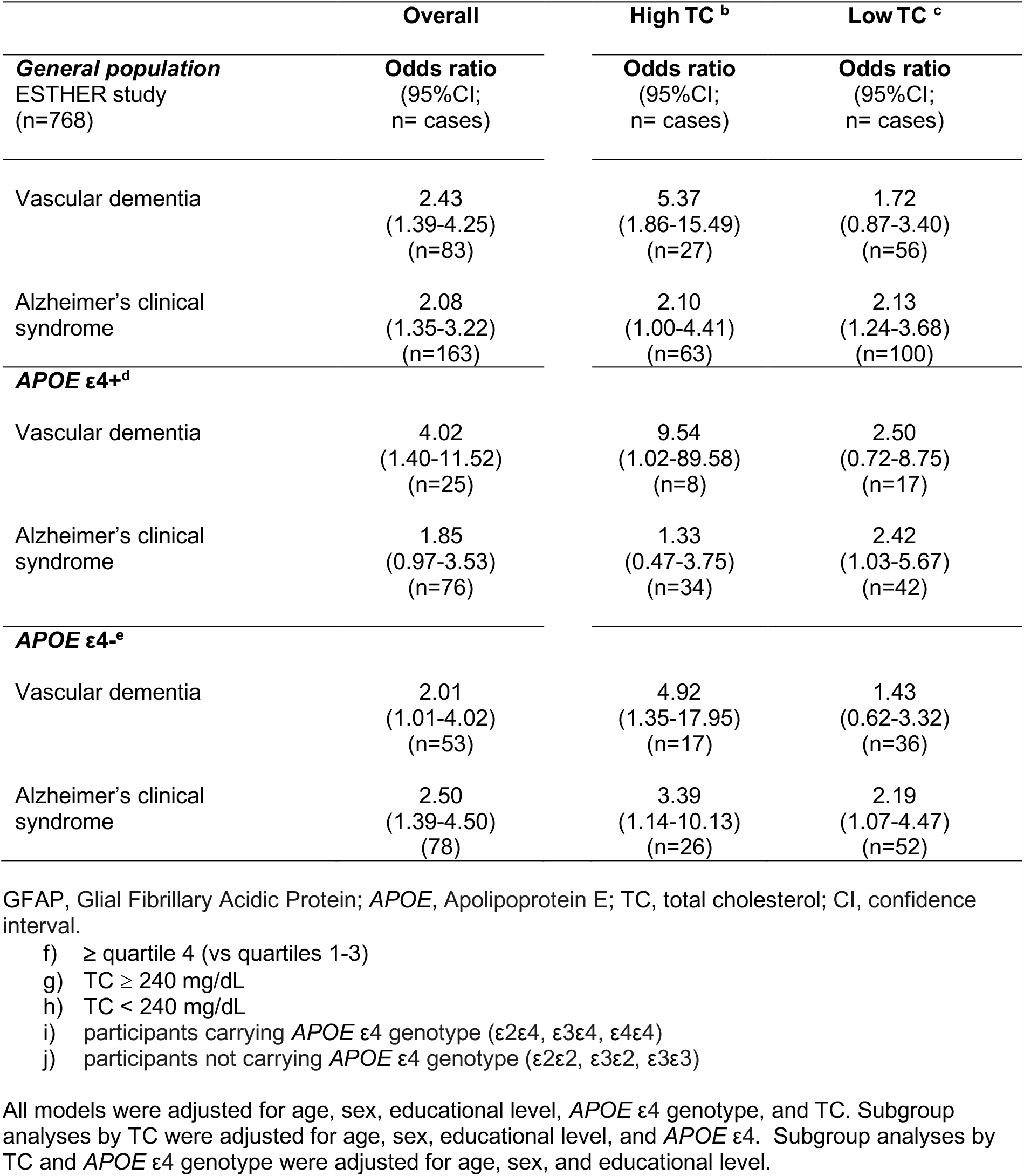
Associations of high^a^ GFAP with vascular dementia and Alzheimer’s clinical syndrome by total cholesterol and *APOE* ε4 status (ESTHER cohort study 2000-2017)

**Supplemental Table 2.** Associations of high ^a^ NfL with vascular dementia and Alzheimer’s clinical syndrome by total cholesterol and *APOE* ε4 status (ESTHER cohort study 2000-2017)

|  | Overall | High TC <sup>b</sup> | Low TC <sup>c</sup> |
| --- | --- | --- | --- |
| **General population**<br>ESTHER study<br>(n=768) | **Odds ratio**<br>(95%CI;<br>n= cases) | **Odds ratio**<br>(95%CI;<br>n= cases) | **Odds ratio**<br>(95%CI;<br>n= cases) |
| Vascular dementia | 1.79<br>(1.03-3.12)<br>(n=83) | 1.95<br>(0.69-5.49)<br>(n=27) | 1.76<br>(0.90-3.43)<br>(n=56) |
| Alzheimer's clinical<br>syndrome | 1.10<br>(0.70-1.74)<br>(n=163) | 1.97<br>(0.92-4.22)<br>(n=63) | 0.80<br>(0.45-1.43)<br>(n=100) |
| **APOE ε4+<sup>d</sup>** |  |  |  |
| Vascular dementia | 0.91<br>(0.31-2.63)<br>(n=25) | 0.81<br>(0.13-4.98)<br>(n=8) | 0.94<br>(0.25-3.58)<br>(n=17) |
| Alzheimer's clinical<br>syndrome | 1.01<br>(0.49-2.09)<br>(n=76) | 2.39<br>(0.77-7.41)<br>(n=34) | 0.56<br>(0.21-1.49)<br>(n=42) |
| **APOE ε4-<sup>e</sup>** |  |  |  |
| Vascular dementia | 2.39<br>(1.24-4.61)<br>(n=53) | 3.20<br>(0.89-11.57)<br>(n=17) | 2.31<br>(1.06-5.05)<br>(n=36) |
| Alzheimer's clinical<br>syndrome | 1.23<br>(0.69-2.19)<br>(n=78) | 1.55<br>(0.55-4.35)<br>(n=26) | 1.03<br>(0.51-2.11)<br>(n=52) |

NfL, Neurofilament light; APOE, Apolipoprotein E; TC, total cholesterol; CI, confidence interval.
a) ≥ quartile 4 (vs quartiles 1-3)
b) TC ≥ 240 mg/dL
c) TC < 240 mg/dL
d) participants carrying APOE ε4 genotype (ε2ε4, ε3ε4, ε4ε4)
e) participants not carrying APOE ε4 genotype (ε2ε2, ε3ε2, ε3ε3)
All models were adjusted for age, sex, educational level, APOE ε4 genotype, and TC. Subgroup analyses by TC were adjusted for age, sex, educational level, and APOE ε4. Subgroup analyses by TC and APOE ε4 genotype were adjusted for age, sex, and educational level.

**Supplemental Table 3.**
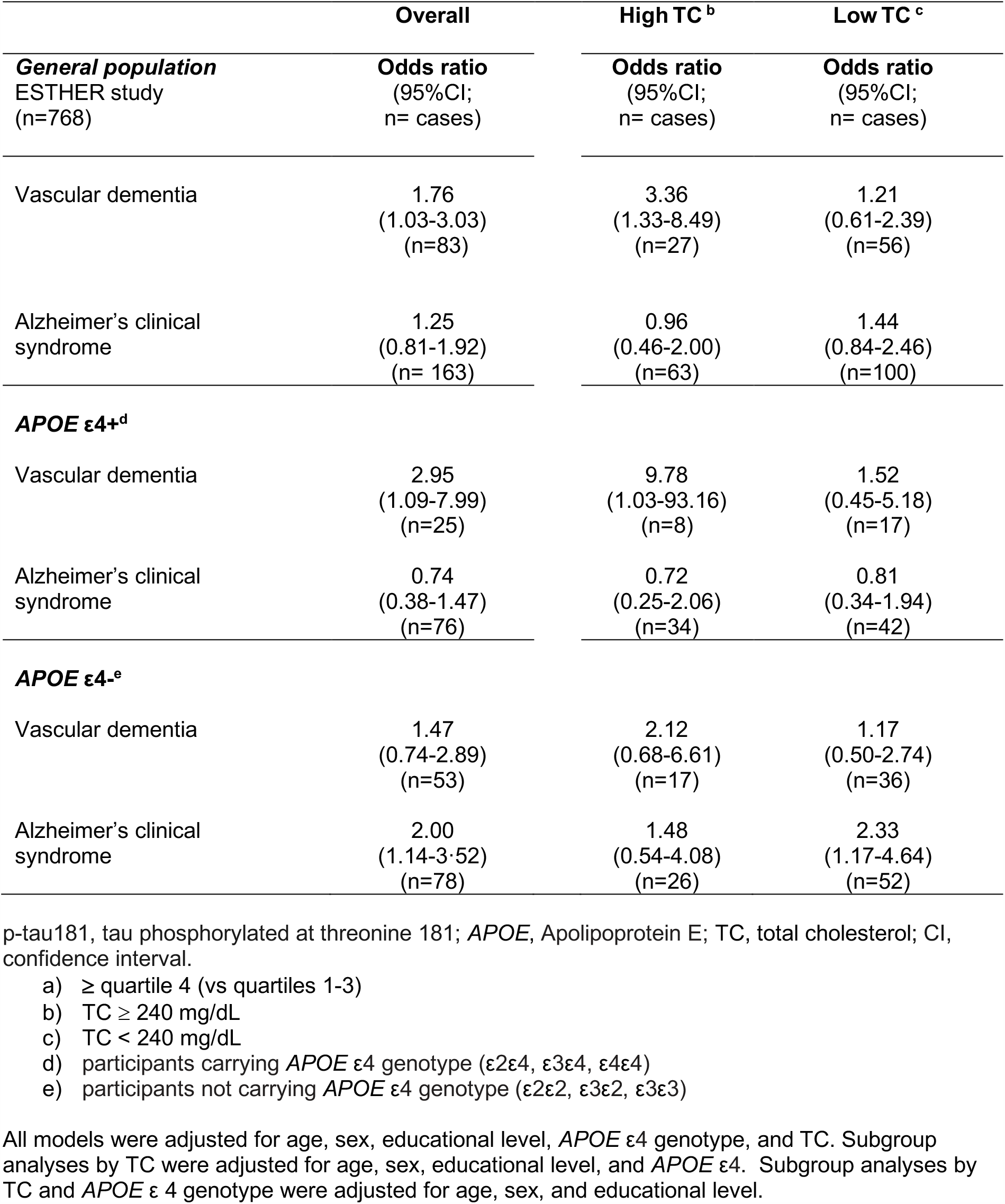
Associations of high ^a^ p-tau181 with vascular dementia and Alzheimer’s clinical syndrome by total cholesterol and *APOE* ε4 status (ESTHER cohort study 2000-2017)

